# Continuity of services for patients with tuberculosis in China in the COVID-19 era

**DOI:** 10.1101/2020.07.16.20150292

**Authors:** Xin Shen, Wei Sha, Chongguang Yang, Qichao Pan, Ted Cohen, Shiming Cheng, Qingshan Cai, Xiaohong Kan, Peilan Zong, Zhong Zeng, Shouyong Tan, Ruixia Liang, Liqiong Bai, Jia’an Xia, Shucai Wu, Peng Sun, Guihui Wu, Cui Cai, Xiaolin Wang, Kaixing Ai, Jianjun Liu, Zheng’an Yuan

## Abstract

It is crucial to maintain continuity of essential services for people affected by tuberculosis (TB). Efforts to deliver these essential services in many global settings have been complicated by the emergence and global spread of SARS-CoV-2 and the pandemic of COVID-19. Understanding how the COVID-19 pandemic has impacted the availability of TB diagnostic and treatment services is critical for identifying policies that can mitigate disruptions of these essential services. China has a dual burden of TB and COVID-19. We conducted a survey and collected data from 13 provinces in China to evaluate the early impact of COVID-19 on TB services and to document interventions that were adopted to maintain the continuity services for TB patients during the pandemic. We use these data to identify additional opportunities which will improve the ability of TB programs to maintain essential services during this crisis. While health systems and underlying epidemiology differ between countries, we believe that sharing China’s experience can inform the design of locally tailored strategies to maintain essential TB services during the COVID-19 pandemic.

## Introduction

The emergence and spread of the novel coronavirus SARS-CoV-2 has caused a pandemic of COVID-19^1^. The COVID-19 pandemic poses unprecedented challenges for health systems. In addition to imposing new demands of these systems to respond to this novel virus, the rapid spread of SARS-CoV2 threatens access to and delivery of essential health services that were needed prior to the pandemic^2^.

Tuberculosis (TB) is the leading infectious cause of death due to a single pathogen. TB diagnosis depends on individuals with symptoms having access to diagnostic facilities, and TB treatment requires daily adherence to antibiotic treatment, often directly observed by healthcare personnel, for six months or longer. TB diagnosis and care is thus a major public health undertaking, and while investment in TB control is one of the single most cost-effective health interventions, TB programs are often operating with limited budgets. Many leading international agencies and organizations coordinating global efforts for TB control, including WHO and STOP-TB Partnership, have voiced concern about the potential of COVID-19 to undermine recent gains in TB control.

New data validate concerns about the detrimental impact of COVID-19 on the operational capacity of TB control programs. A recent modelling study estimated an additional 190, 000 TB deaths (a 13% increase) in 2020 if global TB case detection decreases by an average 25% over a period of three months (as compared to the level of detection before the pandemic), bringing us back to the levels of TB mortality that we had five years ago^3^. Currently, it is urgent to maintain continuity of essential services for people affected by TB during the COVID-19 pandemic.

China faces the dual burden of COVID-19 and TB since first outbreak of COVID-19 in Wuhan in December 2019^4,5^. As COVID-19 emerged in Wuhan, China used aggressive public health interventions to mitigate spread including lockdown of cities, and restriction of public transportation. In addition, there was substantial re-allocation of existing medical resources to meet the needs of the COVID-19 patients. These shifts included the temporary re-designation TB hospitals as special COVID-19 hospitals. Though these measures have thus far succeeded in limiting the local spread and health effects of COVID-19^6^, there are concerns that these efforts may have negative consequences for TB control and care of existing TB patients.

Here we developed a hospital-based survey to better characterize the impact of COVID-19 on TB care in China. We also describe how personnel at TB hospitals and TB programs have developed creative strategies to maintain continuity of care for TB patients during the COVID-19 pandemic. We compare specific metrics of TB diagnosis and treatment during three phases: pre-pandemic phase (2019), the emergency response phase (January to March 2020), and the mitigation phase (April 2020). We also describe anticipated recommendations for next steps. Given the global nature of the COVID-19 crisis, we believe that sharing China’s experience can help other TB programmes minimize disruptions in TB care and TB control during the ongoing struggle against COVID-19.

### Survey Design

The National and Shanghai Anti-tuberculosis Associations organized this survey and invited the representative TB-designated hospitals in provinces which were affected mostly by COVID-19 in China. Overall, thirteen invited TB hospitals from thirteen provinces in different part of the mainland China (including the most affected Wuhan city in Hubei province, Figure 1) participated this questionnaire survey. Each of the included TB hospitals established special hospital-wide crisis teams to evaluate local trends of COVID-19 as well as other diseases, including TB, and centrally coordinate their response to prevent conflicting department-specific strategies. These 13 provinces reported over 50% (414,673) of newly diagnosed pulmonary TB cases in 2018, and reported 91·8% (76,088) of COVID-19 cases until April 30, 2020 in China. Each TB hospital enrolled in this study was within the top three hospitals in terms of province-wide numbers of pulmonary TB in 2018-2019. Although the sample size of the survey is limited, it covers all seven regions in mainland China (Figure 1), and thus the survey results represent the status of tuberculosis diagnosis and treatment services at the provincial level tuberculosis designated hospitals during the COVID-19 outbreak.

**Figure 1.**
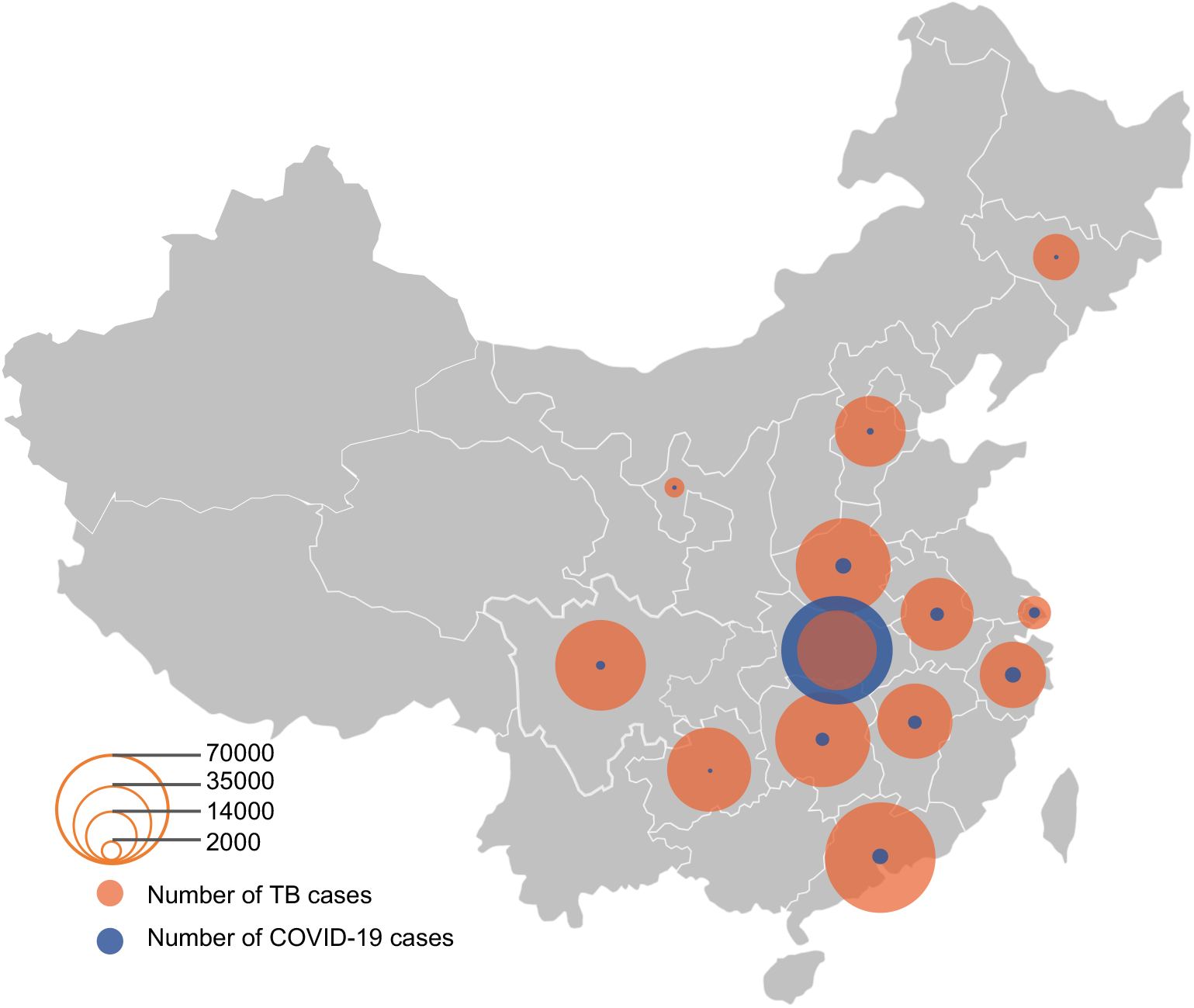
Distribution of annual notified tuberculosis patients in 2018 and COVID-19 patients by April 30, 2020 in the 13 provinces. The size of circle represents the number of tuberculosis (orange) and COVID-19 cases (blue). The Hubei province was the only one with more accumulated COVID-19 cases than the annual notified tuberculosis cases.

In this survey, we divided COVID-19 epidemics in China in three periods: the pre-pandemic phase (2019), the outbreak emergency response phase (January through March, 2020); and the outbreak mitigation phase (April, 2020). We conducted a pilot study in one hospital prior the full scale-up of the survey. We collected information about three aspects of TB diagnosis and care during the outbreak: 1) Changes in hospital-based diagnosis of TB patients; 2) Changes in the availability of hospital-based TB services; and 3) Adjustment of hospital-based TB services.

### Findings

#### Changes in the detection of TB

Compared with the same period of 2019, the number of TB patients diagnosed significantly decreased during the pandemic (Table 1). In the emergency response phase, eleven hospitals reported a median decrease of 25% in the number of TB patients detected. Two hospitals reported small increases in TB diagnoses. In the mitigation phase, ten hospitals had smaller numbers of TB diagnoses compared to April 2019; however, the gap was decreased (median reduction of 15%) compared with the decreases observed in the emergency response phase.

**Table 1.** Impact of COVID-19 on TB services in 13 TB hospitals in China, 2020.

| Category | Pre-pandemic phase | Pandemic phase | Recovery phase |
| --- | --- | --- | --- |
| **TB medical services** |  |  |  |
| Change in No. of TB beds*& | 100% | Decreased<br>(median -75.7%, range -100% ~ -21%) | Decreased<br>(median -43.6%, range -53.3 ~ -5.0%) |
| Change in No. of professional TB staff*& | 100% | Decreased<br>(median -28.5%, range -100% ~ -3%) | Decreased<br>(median -10%, range -38% ~ 0%) |
| No. of hospitals set strict indications for hospitalization | 0 | 38.5% | 23.1% |
| No. of hospitals set restriction for maximum outpatient visits | 23.1% | 30.8% | 23.1% |
| Change in No. of outpatient visits& | 100% | Decreased<br>(median -34%, range -99% ~ -6%) | Decreased<br>(median -20%, range -90% ~ -2%) |
| Change in No. of patients' admission& | 100% | Decreased<br>(median -30%, range -100% ~ -11%) | Decreased<br>(median -18%, range -95% ~ 0%) |
| **Detection of TB patients** |  |  |  |
| Change in No. of TB patients detected & | 100% | Decreased<br>(median -25%, range -100 ~ +11%) | Decreased<br>(median -15%, range -90 ~ +15%) |

\* TB beds and professional TB staff were deployed for COVID-19.
& Compared with the same period of 2019.
Pre-pandemic phase = the same period of 2019, Emergency phase = January – March 2020, Recovery phase = April 2020, TB = tuberculosis.

#### Changes in the availability of hospital-based TB services

In the emergency response phase, nine of the 13 hospitals in our study had been designated as COVID-19 hospitals and shifted at least some fraction of designated (median 75·7%, range, 21·1-100·0%) TB beds for COVID-19 care. Four of the hospitals converted 100% of TB beds to COVID-19 beds during this period. Twelve of the 13 TB hospitals dispatched professional TB staff for COVID-19 service (28·5%, 3·0-100·0%). Among the nine hospitals that continued inpatient services for TB, five (38·5%) set stricter indications for TB hospitalization than had previously been used. For example, only those patients with severe tuberculosis, such as patients with hemoptysis, with massive pleural effusion, or those with drug-resistant TB, would be admitted to the hospital. Ten (77·0%) of 13 hospitals set restrictions for the numbers of outpatient visits. These measures significantly decreased the supply of TB services. Compared with the same period in the previous year (January-March 2019), there were 34% (6-99%) and 30% (11-100%) reductions in the number of outpatient visits and the number of admissions in the emergency response phase, respectively (Table 1).

In the mitigation phase, the fraction of TB medical resources deployed for COVID-19 gradually decreased. Five hospitals still shifted the TB beds for COVID-19 but with a deceased median proportion of 43·6% (5·0-53·3%) compared to emergency response phase. Twelve TB hospitals continue to dispatch professional TB staff for COVID-19, but the proportion of TB staff repurposed for decreased from 28% to 10% (Table 1). The number of hospitals that set stricter indications for hospitalization or set a restriction for maximum outpatient visits both decreased to three (23·1%). Compared with the same period of April 2019, the decline has narrowed for the number of outpatient visits, the number of patients’ admission and discharge for each TB hospitals (Table 1), indicating the gradually restoration of TB medical services. One notable exception is the Jinyintan Hospital in the Wuhan -- the city at the epicenter of this outbreak^7^ -- where the number outpatient visits and inpatient admissions of TB cases was severely reduced (>90%) throughout all COVID-19 outbreak phases.

#### Adjustment of hospital-based TB services

In order to maintain continuity of essential services for people affected by TB during the COVID-19 pandemic, TB hospitals in China made a number of modifications to existing hospital-based TB services. These include changes in treatment monitoring and hospitalization policies, changes to approaches for TB patient support, and changes to infection prevention and control (Table 2).

**Table 2.** Major changes in caring for patients with tuberculosis in China during the COVID-19 pandemic.

| Category | Major changes |
| --- | --- |
| Anti-TB treatment | Provide long-term prescription for TB medications<br>Convert injectable treatments to oral regimens for patients with multidrug-resistant TB<br>Use digital medical technologies for patients' follow-up and clinical assessment |
| Patients' support | Provide web-based education for both TB and COVID-19<br>Develop and disseminate paper educational materials on COVID-19 for patients and their accompany<br>Use digital medical technologies and telephone for patients' counselling |
| Infection prevention and control | Screen all patients and visitors for the symptoms of COVID-19 and fever<br>Ask epidemiological history of COVID-19 for each patient<br>Isolate patients who had symptoms consistent with COVID-19<br>Minimize patients' unnecessary visits to the hospital (online appointment, remote diagnosis and treatment services)<br>Minimize the number of hospitalizations, set strict indications of hospitalization<br>Set restriction for maximum outpatient visits and visitors to accompany<br>All TB patients should be screened for COVID-19 through an appropriate triage system<br>Strengthen environmental disinfection and ventilation system for TB<br>Use upper-room germicidal ultraviolet (UGA) system in consulting room and wards for TB<br>Reduce the usage of usage of bronchoscopy for TB patients<br>All patients and visitors should wear a surgical mask<br>Medical staff is required to wear particulate respirators, the consciousness of wearing particulate respirators is high |

#### Anti-TB treatment

Across the 13 TB hospitals, anti-TB treatment regimens have been adapted for two main reasons: (1) to minimize the number of hospital visits and hospitalizations, and (2) to ensure the continuity of anti-TB treatment. There were five strategies reported by the TB hospitals. First, most hospitals have set stricter indications of hospitalization, as mentioned above. Second, distribution of longer-term prescriptions have been widely used to ensure that TB patients would have sufficient medications for TB treatment. Nine hospitals provided 2-3 months medications, and four hospitals provided 1-month medications for TB patients during the emergency phase, compared to 1-2 week medications prior this pandemic. Third, most hospitals (11 of 13) converted injectable treatments to all oral regimens for patients with multidrug-resistant TB (MDR-TB) to reduce the frequency patient’s visits to hospitals to avoid COVID-19 infection. Fourth, all of the hospitals used digital medical technologies such as instant messaging, and/or telephone calls to follow-up TB patients. Fifth, 11 of 13 hospitals provided drug delivery services for TB patients who started treatment but who were not easily able to visit the hospital to receive medication.

#### Patient support

In our survey, all the hospitals reported that TB patients were often willing to accept suggested preventive measures to avoid infection. Meanwhile, all of 13 hospitals reported that there was increased anxiety among patients with TB compared to the pre-COVID period. The demand for counseling and mental-health assistance is skyrocketing. Questions that patients ask frequently have been typically focused on: 1) the possibility of co-infection with *Mycobacterium. tuberculosis* (*M. tb*) and SARS-Cov-2; 2) how to prevent COVID-19 during clinic/hospital visit, and 3) the impact of COVID-19 on TB treatment and hospital visits.

To address patients’ concerns, TB hospitals have strengthened patient support by adopting multiple interventions. All hospitals had patient-friendly web-based educational material for both TB and COVID-19 and developed and disseminated paper educational materials on both TB and COVID-19. Seven hospitals used instant messaging, short message service and/or telephone to follow up patients and provide consultation. One hospital developed a mobile application for providing personal consultation. Considering the limited in-person interactions with health care workers, and increasing patients’ anxiety caused by COVID-19, patient support appears vitally important to ensure continuity of TB care in the midst of this crisis.

#### Infection prevention and control

The COVID-19 crisis has significantly raised people’s awareness of infection control and personal protection for respiratory infectious diseases^8,9^. All 13 TB hospitals reported that the COVID-19 pandemic resulted in greater general awareness of the importance of infection prevention and control for TB. Consistent with WHO guidelines^10^, hospitals have strengthened the measures on TB infection prevention and control as follows: (1) All TB patients are screened for COVID-19 through an appropriate triage system. TB patients screening negative for COVID-19 can be sent directly to TB services, while TB patients screening positive for COVID-19 are separated within a COVID-19 investigation area. (2) All patients and visitors are required to wear a surgical mask when at the hospitals. (3) All hospitals have strengthened environmental disinfection and ventilation system. Both natural and mechanical ventilation systems are commonly used. (4) Eleven (84·6%) of these 13 TB hospitals use upper-room germicidal ultraviolet (UGA) system in the consulting room and wards for TB. (5) All of the participating hospitals reduced the usage of bronchoscopy for TB patients by a median of 67% (24-100%) during the emergency response phase and by 27·5% (6-90%) in mitigation phase (Table 1).

For general clinical activities, there were several additional measures taken by TB hospitals to prevent transmission. First, all visitors were screened for fever and known exposures to COVID-19. Patients who had symptoms consistent with COVID-19 were isolated rapidly in all 13 hospitals and received urgent diagnostic evaluation. Second, remote medical services were widely implemented for TB care. Eleven TB hospitals provided online appointments to reduce the number of visits and to reduce visit waiting times. The proportion of online appointment increased from 8% (5-90%) in 2019 to 17% (10-100%) during the emergency phase and 19% (12-90%) in mitigation phase. Four hospitals provided online remote diagnosis and treatment services for those who could not visit the hospital. Third, all TB hospitals set stricter restriction for visitors. For patients admitted to the hospitals, three hospitals did not allow any visitors and the other ten hospitals allowed only a single visitor per patient. For patients seeking outpatient care, three hospitals did not allow any visitors to accompany the patient and nine hospitals allowed only a single visitor, the remaining one did not set any restriction.

## Discussion

In this survey, we have gained insight into the impact of the COVID-19 outbreak on the continuity of hospital-based TB services. Most TB services, including diagnosis inpatient and outpatient care, decreased substantially during the COVID-19 emergency response phase. There were two main drivers for these changes: (1) TB hospitals were temporarily converted to designated COVID-19 hospitals to handle the expected pandemic surge; (2) TB hospitals reduced the number of consultations and hospitalizations to reduce the risk of nosocomial transmission of COVID-19.

The observed decrease in TB case detection at the surveyed hospitals raises concerns for TB control efforts. Given the slow secular decline in TB incidence in China^11^, the dramatic decline in TB diagnoses during the pandemic is almost certainly the result of changes in care seeking and access attributable to COVID-19. There were clearly documented limitations in available TB services caused by the response to COVID-19 and other movement restrictions likely led to further barriers for probable TB patients to access diagnosis and care. Further, although our survey does not document this phenomenon, it seems likely that concerns about SARS-CoV-2 transmission in health facilities and on public transportation may have prevented individuals from seeking TB diagnosis or care.

The impact of COVID-19 on the health system and the potential for COVID-19 responses to undermine TB control efforts are already clear. This pandemic presents difficult challenges for national and local TB programs and suggests a need to understand how to ensure that COVID-19 responses disrupt TB services as little as possible. Based on these hospitals’ experiences, we propose two specific recommendations:

### Timely modification of TB services

To mitigate the impact of COVID-19 on TB care and control, national and local TB programs should commit to interventions which can maintain the continuity of essential services for TB patients. For example, provision of adequate stocks of medicines for all TB patients can ensure treatment completion without unnecessary hospital visits to collect medications. As China and many other high TB burden countries remain reliant on in-person and community-based DOTS for TB treatment, modification of current in-person TB services in preference for more innovative patient-centered approaches may improve patients adhere to their treatment during this and future crises. Digital-health technologies like instant messaging, electronic medication monitors, and video-supported therapy^12, 13^ can play an important role in this transition. Online diagnosis and treatment services should be further explored so that patients can have opportunities to access medical care regardless of distance from clinics.

### Integration of services for COVID-19 and TB

There are striking similarities between the COVID-19 and TB. Both can present with respiratory symptoms such as cough, fever, and difficulty breathing. Both are transmitted mainly via close contact. The integration of services for COVID-19 and TB can assist in curbing both epidemics to save lives. Accurate diagnostic tests are essential for both TB and COVID-19. Though tests for the two conditions are different, both should be made available for individuals with respiratory symptoms, so both diseases can utilize the capacity building efforts, along with surveillance and monitoring systems, as well as diagnostic tools such as GeneXpert and chest radiography. In countries with local transmission of COVID-19, the case-finding strategies are being modified to an active approach. Although active TB case finding has not been scale-up in many high TB burden countries, there is a potential for the collaboration between activities of case finding for both TB and COVID-19, such as close contact screening and tracing, testing of patients with severe pneumonia that does not respond to antibiotics, which can help to quickly detect patients with both diseases.

## Conclusions

Our survey reveals a significant disruption of TB diagnosis and care at major hospitals throughout China as a result of COVID-19. These disruptions were most dramatic at the peak of the epidemic, and numbers of TB diagnoses and TB inpatient facilities have gradually returned toward normal levels as COVID-19 incidence receded in April 2020. We also report innovative efforts adopted by hospitals to maintain the continuity of care TB patients during the pandemic. As many countries with high TB burden are still attempting to control COVID-19, our experiences in China offer important lessons for other health systems grappling with maintain TB services in the midst of this crisis. We believe that patient-centered approaches to TB diagnosis and care, which were already being promoted before the emergence of COVID-19, should have new urgency and motivation. We urgently call for additional investment directed toward improving the availability of diagnostics and therapeutics such that equitable access to care can be maintained during times of crisis.

## Contributors

XS, KA, JL, and ZY had the idea for this report, with input from WS, CY, QP, and SC. XS and WS designed the questionnaire. WS, QC, XK, PZ, ZZ, ST, RL, LB, JX, SW, PS, GW, CC, and XW collected the data. XS and CY reviewed published literatures, analyzed the data, made the tables and figure, and wrote the first draft. TC, WS, QP, SC, KA, JL, and ZY reviewed and revised the report. All authors approved the final version.

## Data Availability

The data are available upon request.

## Acknowledgements

This study received funding from the Chinese National Science and Technology Major Projects (grant 2018ZX10715012 to XS and ZY), and the Natural Science Foundation of China (grant 81872679 to XS and QP). CY received funding from the Robert E. Leet and Clara Guthrie Patterson Trust Mentored Research Award. All the funding sources had no role in the preparation of this report.

